# Development and content validation of a questionnaire identifying patients’ functional priorities and abilities after hip or knee arthroplasty

**DOI:** 10.1101/2024.03.20.24304636

**Authors:** Motahareh Karimijashni, Marie Westby, Tim Ramsay, Paul E. Beaulé, Stéphane Poitras

**Affiliations:** School of Rehabilitation Sciences, Faculty of Health Sciences, University of Ottawa, Ottawa, Canada; Clinical Epidemiology Program, Ottawa Hospital Research Institute, Ottawa, Canada; Centre for Aging SMART, Vancouver Coastal Health Research Institute, Vancouver, British Columbia, Canada; School of Epidemiology and Public Health, University of Ottawa, Ottawa, Canada; Division of Orthopaedic Surgery, The Ottawa Hospital, Ottawa, Canada; Faculty of Medicine, University of Ottawa, Ottawa, Canada

**Author notes:** Corresponding Author: Stéphane Poitras, Faculty of Health Sciences, School of Rehabilitation Sciences, University of Ottawa, 200 Lees Avenue (FHS), Ottawa, Ontario K1N 6N5, Canada.

**Keywords:** knee and hip arthroplasty, functional priorities, content validity, questionnaire development

## Abstract

**Background:** To develop a self-report questionnaire evaluating functional priorities after hip or knee arthroplasty and evaluate patients’ understanding of its items and conceptual relevance.

**Methods:** A self-report questionnaire was first developed based on the International Classification of Functioning, Disability, and Health (ICF) core set for osteoarthritis (OA). In the second stage, two research physiotherapists thoroughly reviewed and refined the questionnaire, and another physiotherapist conducted cognitive think-aloud interviews with 18 patients to assess the face and content validity of the questionnaire.

**Results:** All categories and corresponding activities of ICF core set for OA were used to develop the questionnaire. Several questionnaire issues were identified and addressed. Most challenges were related to comprehension, followed by item ordering and visual elements. Patients identified ambiguous wording which we subsequently simplified. Ten activities of the core set were excluded due to lack of face validity, two activities were added, and four activities were modified.

**Conclusion and implication:** The findings suggest that the ICF core set for OA needs to be adjusted for patients undergoing hip or knee arthroplasty and highlight the feasibility of applying a modified core set to assess functional priorities after hip or knee arthroplasty.

## Introduction

Healthcare systems are moving toward patient-centered services by focusing on patient priorities, which are health outcome goals based on what matters most to patients.^1–3^ Patients can contribute to better healthcare quality by sharing their goals and experiences with healthcare providers.^4^ To establish patient-centered management priorities, healthcare providers must understand patient needs and expectations.^5^ Healthcare services tailored to meet patient goals have been demonstrated to enhance outcomes and patient satisfaction while reducing healthcare costs.^6,7^

Hip or knee arthroplasty is an effective procedure for advanced osteoarthritis (OA), with a majority of patients experiencing improved pain, function and health-related quality of life outcomes.^8^ However, there is evidence that 10 to 30% of patients report suboptimal outcomes after hip or knee arthroplasty.^9,10^ Reasons for these poorer outcomes after hip or knee arthroplasty remain unclear.^11^ To define poor outcomes, it is important to understand which aspects of outcomes are important to patients. This knowledge will assist clinicians in identifying patients with suboptimal outcomes, leading to tailored rehabilitation intervention which in turn, contribute to the optimization of results after arthroplasty. Existing outcome measures can be assessed against postoperative patient priorities to determine if they are patient-centered, guiding clinicians to select appropriate measures and assess postoperative outcomes that are important for patients.

Despite the thorough exploration of patients’ expectations and goals before arthroplasty ^12–15^ and their role in the surgical decision-making process,^12,13^ this preoperative data is insufficient for postoperative decision-making. Postoperative information is essential as patients may reassess their goals and expectations after arthroplasty due to the changes in their functional status and perception of health.^16,17^ Many patients have very optimistic preoperative expectations regarding surgery results, potentially leading to unmet expectations and subsequent dissatisfaction.^15,18^ Additionally, only considering preoperative patient expectations makes it challenging to define goals at different time points during the postoperative period.^12,13^

Thus, there is a need to explore postoperative patient priorities after hip or knee arthroplasty, yet limited information exists on this subject. Although few studies^19,20,20^ have identified some aspects of functional priorities at specific postoperative time points, a comprehensive evaluation of patients’ priorities throughout the entire postoperative recovery period is lacking. To address this gap, an instrument is first needed to provide a more detailed insight into patients’ perspectives after surgery. The current study aimed to develop a patient questionnaire to assess functional priorities after hip and knee arthroplasty and evaluate its face and content validity.

## Method

This research was designed following the steps proposed by Burns et al.^21^ and Passmore et al.^22^ and consists of three phases: 1) developing the questionnaire, 2) assessing the face and content validity of the questionnaire,^23^ and 3) pilot testing the draft questionnaire. Figure 1 shows a flow diagram of the methodology for this study. This study was approved by Ottawa Health Sciences Network Research Ethics Board (20230252-01H), and the University of Ottawa Board of Ethics (H-06-23-9425).

**Figure 1.**
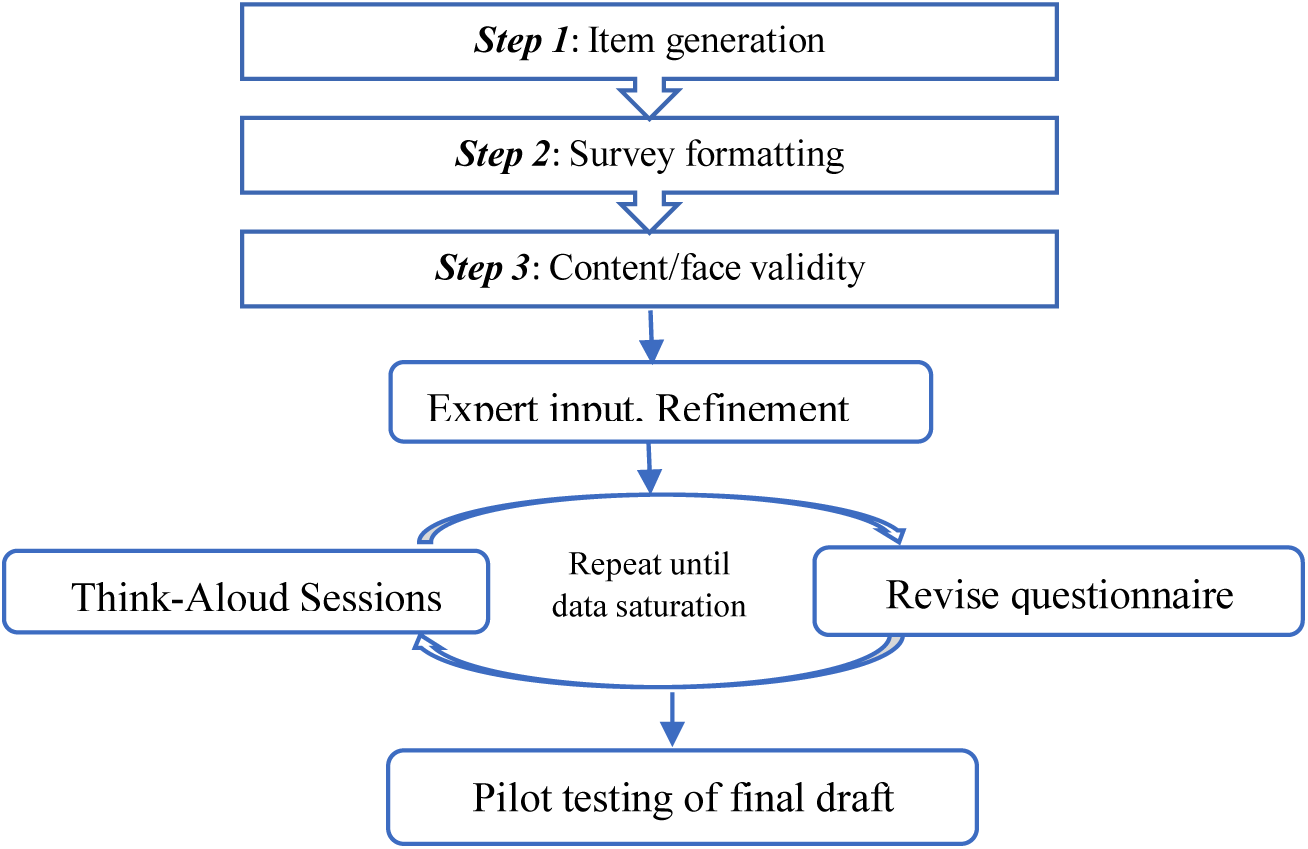
Questionnaire development and cognitive validation.

The development of the questionnaire consisted of three steps:

### Step one: Item generation

The aim of this step is to thoroughly explore potential domains for incorporation into the questionnaire. We selected the International Classification of Functioning, Disability, and Health (ICF) to generate the items in this questionnaire. The ICF is a comprehensive and integrated framework designed to assess the health and health-related states of diverse populations.^24^ Previous research has highlighted the usefulness of ICF as a framework and classification system in the development of more effective questionnaires.^25^ The ICF is organized into two distinct parts: (a) functioning and disability, and (b) contextual factors^24^ (figure 2). The expansive nature of the ICF poses a challenge to its practical application. To address this, lists of relevant categories tailored to certain conditions have been structured into core sets. The ICF core set for patients with OA was developed in 2004 and systematically identifies and categorizes key aspects of functioning and disability related to OA.^26^ It includes 13 chapters for body function (most of which related to the neuromusculoskeletal and movement related functions), 6 chapters for body structure (all of them from the structures related to movement), 19 chapters for activities and participation (mobility, self-care, domestic life, etc.), and 17 for environmental factors (products and technology, support and relationship, etc.). Within these chapters are second-level categories such as changing and maintaining a body position, lifting and carrying objects, walking, going up and down stairs, and moving around. Within each second-level category are third-level activities including lying down, squatting, kneeling, sitting, standing, etc. Figure 3 shows an example of the ICF hierarchically from the activity and participation component. This multifaceted approach provides in-depth insights into the diverse dimensions of daily activities. Only the activity and participation component of the ICF was selected for this questionnaire because enhancing functional outcomes is the main aim of rehabilitation of individuals with OA undergoing arthroplasty to assist them in optimizing their abilities to engage in daily activities at the individual and societal levels.^26,27^

**Figure 2.**
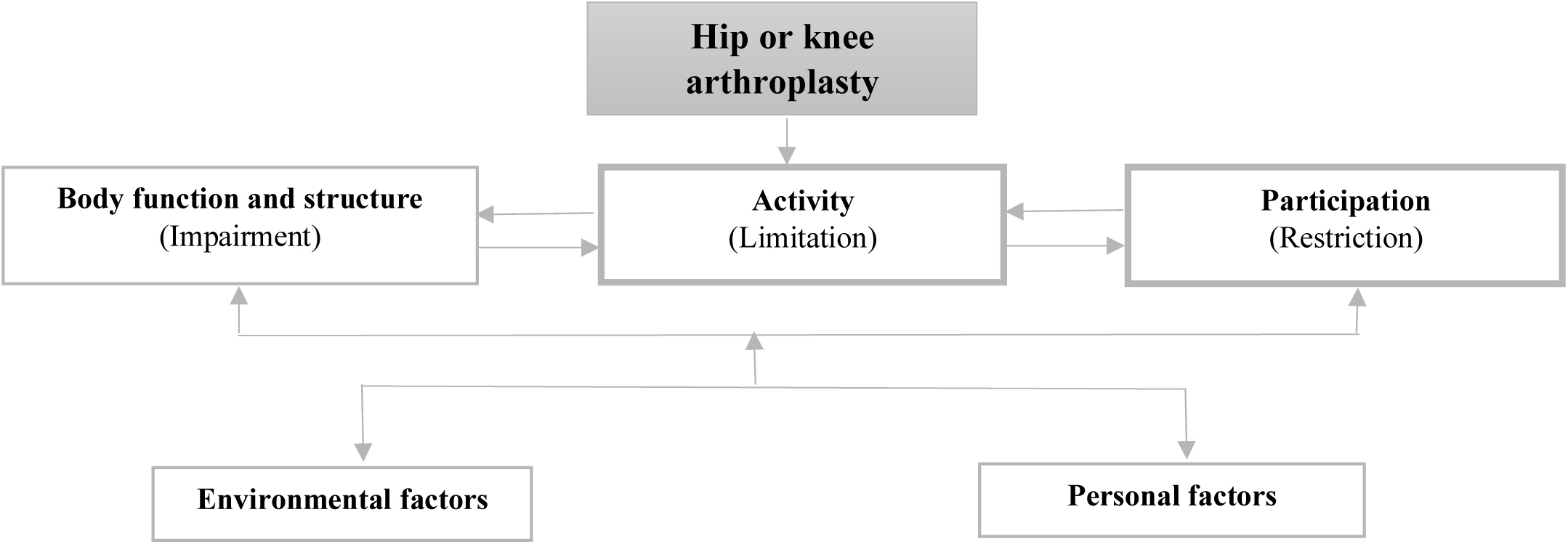
Components of ICF framework.

**Figure 3.**
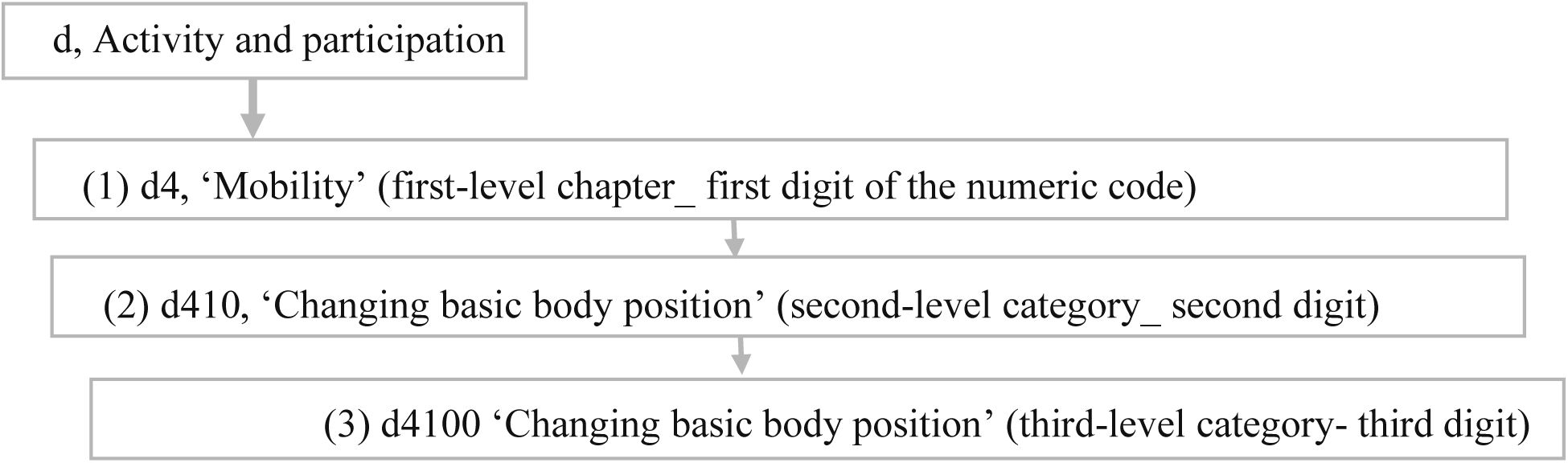
An example of the ICF hierarchically from the activity and participation component.

The OA core set was used to generate items for the questionnaire since previous research has shown the potential clinical utility of this core set in the assessment of arthroplasty outcomes^28^. It encompasses the common range of functional issues faced during the recovery period following hip or knee arthroplasty enhancing the content validity of our questionnaire.^27,28^ Also, it has been validated from both physiotherapists and patient perspectives.^57,60^ The OA core set addresses second-level categories, but the survey included third-level activities to ensure a thorough exploration of the individual’s participation in diverse life activities using original ICF wording. It is worth noting that an ICF core set for total knee arthroplasty was recently developed,^29^ however patients were not involved in the development process.

### Step two: Questionnaire formatting

In this step, the questions and response options were formulated by the first author (MK) with the goal of ensuring conciseness, clarity and interpretability. Each activity is evaluated based on current importance and ability. Participants rate the importance of functional activities on a 4-point scale from “not important” to “very important” and ability to perform activities from “unable to do” to “no difficulty”. A 4-point scale (very important, important, slightly important, and not important)^30^ was used to limit neutrality and force respondents to make a clear directional choice and provide concrete opinions. This scale is also easier to understand, requires less effort to answer ^31^ and maximizes response rates.^32^ We included an image from the ICF illustration library for each activity to enhance comprehension by providing more precise information and facilitating ease in answering.^33^

### Step three: Questionnaire tool validation

#### Validating procedure

The questionnaire went through two stages to assess its face/content validity. First, two physiotherapist research team members (SP, MW) reviewed the questionnaire items and commented on the wording of the questions and determined whether the questions captured the objectives pursued.^34^ Second, a think-aloud process was applied with the intended users of the questionnaire, e.g. patients who had undergone hip or knee arthroplasty. This cognitive interview method assesses validity by investigating response processes.^35^ Patients are prompted to vocalize their thoughts by thinking aloud while answering questionnaire items, allowing them to verbalize thoughts that would typically remain silent during the process.^36^ The think-aloud method allows for investigating the challenges patients may encounter when completing the questionnaire, including issues with comprehension, retrieval, judgment, and responding. This approach aims to ensure the functionality, clarity and user-friendliness of the questionnaire and is thought to give a more realistic picture of the problems compared to more direct interview methods that may interrupt task completion.^37^ The interviewer remains silent so long as individuals continue to think aloud. Participants are consistently encouraged to keep thinking aloud if they stop verbalizing thoughts for more than ten seconds. They are not asked to explain or justify what they are doing nor to report their strategies.

#### Setting and participants

This study was conducted within the orthopedic department at The Ottawa Hospital in Ottawa, Canada, a large academic-affiliated institution situated in an urban setting with a high volume of arthroplasties. The inclusion criteria were adults (18 years or older) who underwent primary elective hip or knee arthroplasty due to OA, had surgery 1, 6, 13, or 26 weeks prior, and who spoke English. Recruitment timing was based on the typical post-arthroplasty recovery pattern.^38,39^ One week post-surgery captured acute recovery. Approximately six weeks post-surgery was selected for the first post-acute measurement as most adverse events usually occur within 30 days of surgery ^40,41^ and most patients can be considered stable six weeks after surgery. The assessments at 13 and 26 weeks were selected to capture the majority of overall functional recovery and reflect the typical timeframe of rehabilitation.^42,43^ Patients with a severe mental impairment, with hearing or speech impairment, and those unable to read were excluded.

#### Sampling and recruitment

We used a phone-based remote recruitment strategy to recruit the patients and purposive sampling. This sampling method allows the recruitment of participants across various criteria, including age, gender, surgery type, post-surgery timepoint. This approach leads to a more representative sample of the whole population and enhances comprehensiveness of the study.^44,45^ For this purpose, screening for potential patients was first performed via Electronic Health Record (EPIC) at the Ottawa Hospital which contains comprehensive information regarding patient’s medical history. After identifying eligible participants, we then manually reviewed charts for study confirmation. One researcher (MK) then phoned individuals with documented consent for research in their EPIC chart. Consent for this study was obtained verbally over the phone or in written format.

#### Data collection

Consenting participants attended a single, cognitive interview session. Think-aloud interviews were held in person or virtually using Microsoft Teams. All interviews were carried out by one interviewer (MK) who is an experienced physiotherapist and a PhD candidate in rehabilitation sciences. After explaining the think-aloud process, participants were asked whether they found the concepts and items of the questionnaire to be relevant, appropriate, and comprehensible in accordance with the questionnaire objectives. After each interview, participants were encouraged to share their general comments on the questionnaire. All sessions were audio-recorded using an encrypted digital audio recorder and subsequently transcribed for analysis. The interviewer collected the demographic data, including age, sex, gender, body mass index, surgery date, and time since surgery. Protection of privacy and confidentiality of participants was ensured by de-identifying data using their unique study IDs. All data, including master log, audio recordings, transcribed recordings, and consent forms were stored in a password-protected shared folder which could only be accessed by the research team. Data security was also ensured with physical safeguards by storing the hard copy of transcribed interviews and data analysis in a locked cabinet and research office in the orthopedic division at The Ottawa Hospital.

#### Sample Size

Recruitment of participants was continued until data saturation,^46^ when no new problems arose for two consecutive participants. For this purpose, the analysis of each transcript was carried out iteratively after its corresponding interview to monitor for data saturation.

#### Data analysis

Data was entered into IBM SPSS (version 28) to produce descriptive statistics of participant demographics. One researcher (MK) analyzed interview transcripts using the inductive content analysis approach (50) and regularly debriefed with other team researchers (SP, MW) to discuss problems raised and modify questionnaire items.^49^

## Results

We developed our questionnaire based on the ICF core set for OA, and validated it through two stages, including expert refinement, and cognitive think-aloud interview. See Appendix A for the final version of the questionnaire.

## Initial questionnaire development by the research team

All third-level activities of the OA core set were critically evaluated by research team members, including three experienced physiotherapists (SP, MW, MK), with some items being removed or revised when reaching a consensus (table 2).

### Removed items

Nine activities were removed: menstrual care (d5302) was excluded to ensure the questionnaire’s applicability to all genders; choosing appropriate clothing (d5404), maintaining head position (d4155), fine hand use (d440), hand and arm use (d445) since they do not involve lower extremities; carrying on the head (d4304), human-powered vehicles (d4700), using humans for transportation (d4703), driving animal-powered vehicles (d4752) since they are less applicable in developed countries (table 2).

### Revised items

Only the second-level categories of “toileting” (d530), “acquisition of goods and services” (d620), “doing housework” (d640), “assisting others” (d660), “intimate relationship” (d770), and “community life” (d970) were used as the details provided by the third level activities are less relevant for functional assessment after knee or hip arthroplasty.^29^ Since the abilities related to hobbies, play, crafts, arts and culture were similar, they were merged into the category “Hobbies/ play /Crafts/ Arts and culture”

“Lying down” (d4100) was split into lying down and getting up. “Center of gravity” was removed from “shifting the body’s center of gravity” (d4106) to simplify the term. To highlight the movement of the activity, “squatting” (d4101), “kneeling” (d4102), “sitting” (d4103) and “standing” (d4104) were modified to “squatting down”, “kneeling down”, “sitting on a chair” and “rising from a chair”, respectively. To improve clarity, “using private motorized transportation” (d4701), “using public motorized transportation”(d4702), “driving human-powered transportation” (d4750), and “driving motorized vehicles” (d4751) were changed to “getting in and out of a car”, “using public transportation”, “biking”, and “driving” respectively. Examples of “slope, uneven, or moving surfaces, such as grass, gravel and snow” and “such as walking around a marketplace or shop, around or through crowded areas” were added to “walking on different surfaces” (d4501), and “walking around obstacles” (d4503) from the ICF description. “Climbing” (4551) was divided into climbing, and going up and down stairs based on the updated 2018 version of the ICF.^50^ “Drying oneself” (d5102) was changed to drying your body and the explanation of “drying some part or parts of your body or the whole body, such as after washing” was added based on the ICF description. “Acquisition of goods and services” (620) was changed to shopping. The explanation of “cleaning the house, washing clothes, using household appliances, storing food and disposing of garbage” and “assisting others with their learning, communicating, self-care, and movement, within the house or outside” were added to “doing housework” (d640) and “assisting others” (d660) respectively from the ICF description (table 2).

We modified several activity illustrations to make them more relevant to developed countries or the lower limb: maintaining a lying position was changed from prone to supine; sitting was changed from the floor to a chair; lifting was changed from an object on a table to the floor; carrying in the hand was changed from a glass to a suitcase; carrying on shoulders, and back was changed from a child to a backpack; putting on clothes was change from upper extremity to lower extremity (table 2).

## Think-aloud interviews

Eighteen participants were recruited in the study. Detailed participant demographic information is presented in Table 1. Three in-person and 15 virtual interviews were conducted between July and September 2023. Each interview lasted between 30 minutes and one hour.

**Table 1.** Think-aloud interview demographic characteristics.

|  |  |  |
| --- | --- | --- |
| **Age, yrs.**<br>**Median (range)** |  | 63 (51-76) |
| **Sex, No.** | Female | 10 |
|  | Male | 8 |
| **Gender, No.** | Woman | 10 |
|  | Man | 8 |
| **BMI, kg/m<sup>2</sup>**<br>**Median (range)** |  | 31 (19.18-52.34) |
| **Surgery Type,**<br>**No.** | Total knee arthroplasty | 10 |
|  | Total hip arthroplasty | 8 |
|  | Hip resurfacing | 1 |
| **Time after**<br>**Surgery, No.** | 2 weeks postop | 6 |
|  | 6 weeks postop | 3 |
|  | 13 weeks postop | 5 |
|  | 26 weeks postop | 4 |

**BMI:** Body mass index; **No.:** Number; **yr:** year

## Issues identified and corresponding modifications

The issues faced by participants were classified into three themes: clarity/comprehension, item order, and visual elements. All participants conveyed that they did not find the number of questions burdensome or overwhelming. Table 2 shows participant quotes from the think-aloud interviews. In the following section, corresponding quotes are referenced as “*Quote X*”.

**Table 2.** OA ICF core set categories revision with verbatim quotations from participants during the think aloud interview.

| ICF level | Original activity and participation of ICF core set for OA | Decision | Reason for removal or revision | Modified version |
| --- | --- | --- | --- | --- |
| First | **d4 Mobility** |  |  |  |
| Second | **d410 Changing basic body position** | Remove | Using third level. | NA |
| Third | d4100 Lying down | Revise | <u>Research group consensus:</u> "getting up" was added to match the ICF description.<br><u>Think-aloud:</u> "In bed" was added.<br>**Quote 1:** "That question is not clear like if it just asked me lying down in bed, I would say no difficulty. It's not really. I never had difficulty with, but on the floor getting down and getting up off the floor is umm the challenge and that's an important thing for me. But the question doesn't talk about those. all depends on the height of the of the horizontal service surface that you want there. " | Lying down in bed and getting up |
|  | d4101 Squatting | Revise | <u>Research group consensus:</u> "down" was added to highlight the movement of the activity.<br><u>Think-aloud:</u> "partial or full squat" was added.<br>**Quote 2:** "It depends on how deep the squat goes, like I can't do a full squat, but I can definitely sit in a chair and get up and do that. So that question's kind of a little ambiguous." | Squatting down (partial or full squat) |
|  | d4102 Kneeling | Revise | <u>Research group consensus:</u> "down" was added to highlight the movement of the activity.<br><u>Think-aloud:</u> "on the ground" was added.<br>**Quote 3:** "again it depends on the level, right? If I'm going to kneel down to the floor, that's one thing. If I'm going on to a Uh, bench or something? That's a different thing. UM. I'm assuming you're kneeling on the floor." | Kneeling down on the ground |
|  | d4103 Sitting | Revise | <u>Research group consensus:</u> "on a chair" was added to highlight the movement of the activity.<br><u>Think-aloud:</u> "average height" was added.<br>**Quote 4:** "It kind of depends on the chair. I avoid squishy ones because they're hard to get out of it, but I would say no difficulty. It's average chair"<br><u>Think-aloud:</u> " with or without arms" was added.<br>**Quote 5:** "this is difficult because some chairs have arms. Some chairs don't, so a chair without arms is a lot harder to get out of than a chair with arms, right? Because I can use the arms to get out of the chair. So that's again confusing. Again, you should be telling you asking what kind of chair with arms. Chair without arms because the dining room chair has no arm" | Sitting on a chair (average height with or without arms) |
|  | d4104 Standing | Revise | <u>Research group consensus:</u> Modified to "rising from a chair" to highlight the movement of the activity.<br><u>Think-aloud:</u> "average height" was added. | Rising from a chair (average height with or without arms) |
|  |  |  | **Quote 6:** "A chair can be many things and very different in different situations, so I don't know"<br><u>Think-aloud:</u> "with or without arms" was added.<br>**Quote7:** "It all depends on what some people are used or like.<br>So it'd be nicer to have some kind of clarification there, like a hard chair or a or a dining room chair or something like that, or change it all" |  |
|  | d4105 Bending | Revise | <u>Think-aloud:</u> "over or to the side (such as reaching down for an object)" was added.<br>**Quote 8:** "What kind of bend are you asking for? Are you talking like a bend from the back or a bend from the knees?" | Bending over or to the side (such as reaching down for an object) |
|  | d4106 Shifting the body's centre of gravity | Revise | <u>Research group consensus:</u> "Center of gravity" was removed to simplify.<br><u>Think-aloud:</u> "From side to side" was added with the explanation "such as, moving from one foot to another while standing or shifting your weight while sitting"<br>**Quote 9:** "It would be better if you were clear on the question, like if it meant shifting your body weight to one leg like you're the knee that you have replaced, that would be a better question, or if you could word it around that" | Shifting the body from side to side (such as moving from one foot to another while standing or shifting your weight while sitting) |
|  | d4107 Rolling over | Revise | <u>Think-aloud:</u> "Such as back to side" was added.<br>**Quote 10:** "You wanna roll over something to think about? Is this just this is when you're sleeping because the picture shows the person on the bed, right? So yeah, maybe you could say for side sleepers or how however you want to do it. I mean umm, because I know with me when I'm sleeping at night, it's a little more difficult because I still have a pillow. I'm a side sleeper. I still have a pillow between my legs." | Rolling over (such as back to side) |
| **Second** | **d415 Maintaining a body position (Second-level category)** | Remove | Using third level. | NA |
| **Third** | d4150 Maintaining a lying position | Revise | <u>Think-aloud:</u> "maintaining" was replaced with "holding" to highlight the static posture of the activity<br>**Quote 11:** "I think that this is somewhat at the very least somewhat repetition in my mind that so unless you want to be more clear if presenting the question, uh, to to make it a little more evident or obvious to people such as myself trying to respond what you mean by as what would distinguish between the two? "<br><u>Think-aloud:</u> "during the day" was added.<br>**Quote 12:** "I don't lie down during the day unless I'm napping which I am not doing a lot."<br>**Quote 13:** "So how do you want me to answer that? That's so that's the way I sleep. I guess more than one hour." | Hold a lying position during the day |
|  | d4151 Maintaining a squatting position | Revise | <u>Think-aloud:</u> "maintaining" was replaced with "holding" to highlight the static posture of the activity. See **quote 11**.<br><u>Think-aloud:</u> The term "partial or full squat" was added.<br>**Quote 14:** "I can squat, but I cannot do a deep squat where my quadriceps are parallel to the ground like I can only squat like I can squat this much. So how many degrees is that? I don't know what is that you tell" | Hold a squatting position (partial or full squat) |
|  | d4152 Maintaining a kneeling position | Revise | <u>Think-aloud</u>: The "maintaining" was replaced with "holding" to highlight the static posture of the activity. See **quote11**. | Hold a kneeling position |
|  | d4153 Maintaining a sitting position | Revise | <u>Think-aloud</u>: The "maintaining" was replaced with "holding" to highlight the static posture of the activity. See **quote 11**. | Hold a sitting position |
|  | d4154 Maintaining a standing position | Revise | <u>Think-aloud</u>: The term "maintaining" was replaced with "holding" to highlight the static posture of the activity. See **quote 11**.<br><u>Think-aloud</u>: "such as standing at the sink or in a line" was added to highlight the static movement of the activity<br>**Quote 14**: *I'm thinking of if I go out with friend and we're going someplace, you know, we can easily walk around for more than an hour. So is that correct? OK, so if I'm standing, say at the sink, Something like that.* | Hold a standing position (such as standing at the sink or in a line) |
|  | d4155 Maintaining head position | Remove | <u>Research group consensus</u>: Does not involve lower extremity activities | NA |
| **Second** | **d430 Lifting and carrying objects** | Remove | Using third level. | NA |
| **Third** | d4300 Lifting | Revise/<br>add | <u>Think-aloud</u>: "From the floor" was added and divided into light and heavy objects.<br>**Quote 15**: *"I think for me it looks a little bit heavy and it's on the floor which is very different from lifting a grocery bag off the table".*<br><u>Think-aloud</u>: "Less than 1 pound" was used for light and "more than 10 pounds" for heavy.<br>**Quote 16**: *"I lift an object. No difficulty. Lift a heavy object. I couldn't. I wouldn't even try, so the objects I would try to lift an average weight I don't have any clue what an average weight is. I have no idea. Are you talking about a 10-pound bag of potatoes and trying to lift a 10-pound bag of potatoes, or are you talking about my pillow that fell off the bed and I'm trying to lift it and put it back on? Umm, I don't know."* | Lifting a light object from the floor (less than 1 pound)<br>Lifting a heavy object from the floor (more than 10 pounds) |
|  | d4301 Carrying in the hands | Revise | <u>Think-aloud</u>: "such as a suitcase or grocery bag" was added.<br>**Quote 17**: *"Are we talking about, like the suitcase? Are we talking about carrying like this? You're carrying like this like grocery bags. Umm, are different than like carrying laundry baskets then I don't know, but how specifically would be?"* | Carrying in the hands (such as a grocery bag or suitcase) |
|  | d4302 Carrying in the arms | Retain | NA | Carrying in the arms |
|  | d4303 Carrying on shoulders, hip and back | Revise | <u>Think-aloud</u>: "hip" was omitted.<br>**Quote 18**: *"For the hip, I would, but don't know, never do this, but I just don't carry it like that. Umm for the other two I have no difficulty for this shoulder and back"*<br><u>Think-aloud</u>: "such as a backpack" added.<br>**Quote 19**: *"you might want to give an example for that. Such as like you looks like a picture of a backpack or what would you carry on your shoulder was like the young child."* | Carrying on shoulders or back (such as a backpack) |
|  | d4304 Carrying on the head | Remove | <u>Research group consensus</u>: Less applicable in developed countries | NA |
|  | d4305 Putting down objects | Revise | <u>Think-aloud</u>: "On the floor" was added. | Putting down objects on the floor |
|  |  |  | **Quote 20:** "Would this be like somebody handed you something and then you have to put it down or like putting it down on a table as opposed to putting it down on the floor. So you don't wanna clarify how you're putting it down because it's for me, there's a difference between just bending over, putting it down, and then kneeling, putting it down" |  |
| **Second** | **d440 Fine hand use** | Remove | <u>Research group consensus:</u> Does not involve lower extremity activities | NA |
| **Second** | **d445 Hand and arm use** |  | <u>Research group consensus:</u> Does not involve lower extremity activities | NA |
| **Second** | **d450 Walking** | Remove | Using third level. | NA |
| **Third** | d4500 Walking short distances | Revise | <u>Think-aloud:</u> "less than 1 km" was added.<br>**Quote 21:** "how far can you walk? Can you walk 100 meters? Can you walk 200 meters? And you walk 2 kilometers. Can you walk 10 kilometers?"<br>**Quote 22:** "Don't think I can do a kilometer yet. I don't think I can do that yet. I don't know how many steps are in a km. I do everything by steps right? It was 850 steps that I could do in December was 450. I know I'm up. I'm almost at my 800 again, but I don't think that's a kilometer."<br><u>Think-aloud:</u> "On flat surfaces" was added.<br>**Quote 23:** Flat surface or hills doesn't matter to you. Then you should say that, you don't say that. So it like, so you don't care about steps or stairs or anything like that in this one, right. You want to be flat surfaces, so no stairs, no hills. You should be saying that. | Walking short distances (less than 1km) on flat surfaces |
|  | d4501 Walking long distances | Revise | <u>Think-aloud:</u> "more than 1 km" and "On flat surfaces" were added. See **quotes 21-23** | Walking long distances (more than 1km) on flat surfaces |
|  | d4502 Walking on different surfaces | Revise | <u>Research group consensus:</u> Examples of "slope, uneven, or moving surfaces, such as grass, gravel and snow" were added from the ICF description.<br><u>Think-aloud:</u> "ice" as an example was removed.<br>**Quote 24:** "walking on ice even if I didn't have a hip problem, I hate it. So, I would say with a lot of difficulty. So, I'd say it depends on which of these you know, because that's quite a range of surfaces. So, walking on grass sloping all that no problem ice there would be difficulty." | Walking on different surfaces (slope, uneven, or moving surfaces, such as grass, gravel and snow) |
|  | d4503 Walking around obstacles | Revise | <u>Research group consensus:</u> "such as walking around a marketplace or shop, around or through crowded areas" was added. | Walking around obstacles (such as walking around a marketplace or shop, around or through crowded areas) |
| **Second** | **d455 Moving around** | Remove | Using third level. |  |
| **Third** | d4550 Crawling | Remove | <u>Think-aloud:</u> lack of face validity for patients after arthroplasty<br>**Quote 25:** "How is important to be able to crawl right? Is this really necessary? Like I don't know how many people crawl unless we're in the military or drunk. Did you need that though? Really to know if a person can crawl." | NA |
|  | d4551 Climbing | Revise/add | <u>Research group consensus:</u> "going up and down stairs" category (d451 going up and down stairs) was added. | Going up stairs (10 to 12 stairs) |
|  |  |  | <u>Think-aloud</u>: "ladder" and "rocks" were removed.<br>**Quote 26**: *I'm not sure I would have that in the same question. Maybe I would have that as two questions. Well, how different would be to climb on rocks? How difficult it would be to climb a ladder or climb stairs or climb curves? Climb the ladder is different than climbing rocks because rocks are uneven.*<br><u>Think-aloud</u>: going up and down stairs was divided into two separate questions, since patients noted that the ability to ascend stairs differs from the ability to descend.<br>**Quote 27**: *"Did you want to break that down? Because I know with knee surgery and knees there's a big difference between people who can go upstairs good because downstairs is with pain. So wouldn't you ought to be more specific so that you have you can figure out where they're pains being generated from either going up or down the stair."*<br><u>Think-aloud</u>: "10 to 12 stairs" were added.<br>**Quote 28**: *"it's not important at this point, or I'll say slightly if you I do have two little steps at the front of the house, so I can do it, but so I'm going to say slightly important because I don't go, I'm going to start going out more, but these are two little steps and I can do it with a little difficulty."* | Going down stairs (10 to 12 stairs)<br>Going up a curb or step |
|  | d4552 Running | Revise | <u>Think-aloud</u>: "jogging" was incorporated within the running activity<br>**Quote 29**: *"what do you mean by running? Is jogging running? Joggers aren't runners. You could probably just put both jogger slash running and just cover both bases because all you're interested is the movement cause somebody who jogs can jog faster than some people can run, but there is a difference, right? So I'd include both."* | Running/jogging |
|  | d4553 Jumping | Retain | NA | Jumping |
|  | d4554 Swimming | Remove | <u>Think-aloud</u>: "swimming" was moved from the "moving around" section to a sports example.<br>**Quote 30**: *"the only thing that I engage in sports would be swimming. And that's not really a group or anything activity really. Do you consider swimming as a sport?"* | NA |
| **Second** | **d470 Using transportation** | Remove | Using third level. |  |
| **Third** | d4700 Using human-powered vehicles | Remove | <u>Research group consensus</u>: Less applicable in developed countries | NA |
|  | d4701 Using private motorized transportation | Revise | <u>Research group consensus</u>: Changed to "getting in and out of a car"<br><u>Think-aloud</u>: "such as own car or taxi" was added.<br>**Quote 31**: *"Maybe you put in brackets (taxi, friend's car), underneath the main question, you specify the parameters."*<br>**Quote 32**: *"Umm no private. Ohh no look. OK, this is a taxi. So they're calling that private. OK. So, whether so public? Yeah, it's a separate question. Getting on a bus or a train, getting in a taxi."* | Getting in and out of a car (such as own car or taxi) |
|  | d4702 Using public motorized transportation | Revise | <u>Research group consensus</u>: Changed to "using public transportation"<br><u>Think-aloud</u>: "such as a bus or train" was added. See **quotes 31 and 32**. | using public transportation (such as a bus or train) |
|  | d4703 Using humans for transportation | Remove | <u>Research group consensus</u>: Less applicable in developed countries | NA |
| **Second** | **d475 Driving** | Remove | Using third level. |  |
| **Third** | d4750 Driving human-powered transportation | Revise | <u>Think-aloud</u>: Changed to "biking"<br>**Quote 33**: "Umm, without the picture here. What? What are you talking about?"<br><u>Think-aloud</u>: "to go to a place or recreational" was added<br>**Quote 34**: "what happens if I just want to do recreational biking? So wouldn't that be redundant? Again, you're worried about transportation." | Biking (to go to a place or recreational) |
|  | d4751 Driving motorized vehicles | Revise | <u>Research group consensus</u>: Changed to "driving vehicles" | Driving vehicles |
|  | d4752 Driving animal-powered vehicles | Remove | <u>Research group consensus</u>: Less applicable in developed countries | NA |
| **First** | **d5 Self-care** |  |  |  |
| **Second** | **d510 Washing oneself** | Remove | Using third level. | NA |
| **Third** | d5100 Washing body parts | Revise | <u>Think-aloud</u>: "lower" and "such as legs and feet" added.<br>**Quote 35**: "Are these questions only for a hip and knee replacement, or what is it for? OK, so I can't imagine anyone having problems washing their hands, face or hair or nails because of this." | Washing lower body parts (such as legs and feet) |
|  | d5101 Washing whole body | Retain | NA | Washing whole body |
|  | d5102 Drying oneself | Revise | <u>Research group consensus</u>: Changed to "drying your body" and added "drying some part or parts of your body or the whole body, such as after washing" based on the ICF description. | Drying your body (drying some part or parts of your body or the whole body, such as after washing) |
| **Second** | **d530 Toileting (Second-level category)** | Retain | <u>Think-aloud</u>: Changed to "going to the toilet" and added "such as getting to the washroom, getting into and out of position, manipulating clothing, and cleaning oneself" based on the ICF description.<br>**Quote 36**: "Do they mean doing the discharge or do they mean just getting to the toilet? I guess it would be getting to the bathroom would be. Is that what they're meaning?" | Going to the toilet (such as getting to the washroom, getting into and out of position, manipulating clothing, and cleaning oneself_ |
| **Third** | d5300 Regulating urination | Remove | <u>Research group consensus</u>: Using second level. | NA |
|  | d5301 Regulating defecation | Remove | <u>Research group consensus</u>: Using second level. | NA |
|  | d5302 Menstrual care | Remove | <u>Research group consensus</u>: Using second level. | NA |
| **Second** | **d540 Dressing** | Remove | Using third level. | NA |
| **Third** | d5400 Putting on clothes | Retain | NA | Putting on clothes |
|  | d5401 Taking off clothes | Retain | NA | Taking off clothes |
|  | d5402 Putting on footwear | Revise | <u>Think-aloud</u>: Added the example of "shoes or socks"<br>**Quote 37**: "Maybe you could put shoes and socks in brackets for question three and four." | Putting on footwear (shoes or socks) |
|  | d5403 Taking off footwear | Revise | <u>Think-aloud</u>: Added the example of "shoes or socks"<br>**See quote 37.** | Taking off footwear (shoes or socks) |
|  | d5404 Choosing appropriate clothing | Remove | <u>Research group consensus</u>: Does not involve lower extremity activities | NA |
| **First** | **d6 Domestic life** |  |  |  |
| **Second** | **d620 Acquisition of goods and services** | Revise | <u>Research group consensus:</u> Changed to "shopping"<br>Think-aloud: Changed to "going shopping"<br>**Quote 38:** "it's just any kind of shopping, right? Doesn't matter if it's clothes or whatever; it's just shopping. Really shopping so? Do you care? Do you wanna elaborate on that? Because I can do online shopping. Do you care? Because it's online shopping or I'm shopping at, getting out and shopping. I know, but that's your, you said shopping." | Going shopping |
| **Third** | d6200 Shopping | Remove | <u>Research group consensus:</u> Using second level. | NA |
|  | d6201 Gathering daily necessities | Remove | <u>Research group consensus:</u> Using second level. | NA |
| **Second** | **d640 Doing housework** | Revise | <u>Research group consensus:</u> Added the explanation of "cleaning the house, washing clothes, using household appliances, storing food and disposing of garbage" from the ICF description. | Doing housework (cleaning the house, washing clothes, using household appliances, storing food and disposing of garbage) |
| **Third** | d6400 Washing and drying clothes and garments | Remove | <u>Research group consensus:</u> Using second level | NA |
|  | d6401 Cleaning cooking area and utensils | Remove | <u>Research group consensus:</u> Using second level. | NA |
|  | d6402 Cleaning living area | Remove | <u>Research group consensus:</u> Using second level. | NA |
|  | d6403 Using household appliances | Remove | <u>Research group consensus:</u> Using second level. | NA |
|  | d6404 Storing daily necessities | Remove | <u>Research group consensus:</u> Using second level. | NA |
|  | d6405 Disposing of garbage | Remove | <u>Research group consensus:</u> Using second level. | NA |
| **Second** | **d660 Assisting others** | Retain | <u>Research group consensus:</u> the explanation of "assisting others with their learning, communicating, self-care, and movement, within the house or outside" from the ICF description.<br><u>Think-aloud:</u> "with their learning, communicating" in the example was excluded.<br>**Quote 39:** "It's assisting others, with their learning, communicating self-care and movement is kind of a little different than assisting people in general" | Assisting others (assisting others with their self-care, movement, within the house or outside) |
| **Third** | d6600 Assisting others with self-care | Remove | <u>Research group consensus:</u> Using second level. | NA |
|  | d6601 Assisting others in movement | Remove | <u>Research group consensus:</u> Using second level. | NA |
|  | d6602 Assisting others in communication | Remove | <u>Research group consensus:</u> Using second level. | NA |
|  | d6603 Assisting others in interpersonal relations | Remove | <u>Research group consensus</u>: Using second level. | NA |
|  | d6605 Assisting others in health maintenance | Remove | <u>Research group consensus</u>: Using second level. | NA |
| **Second** | **d770 Intimate relationships** | Revise | <u>Think-aloud</u>: Changed to "Sexual relationships"<br>**Quote 40**: "it depends on what you mean by intimate. Now when you say intimate, are you just talking sex intercourse?" | Sexual relationships |
| **Third** | d7700 Romantic relationships | Remove | <u>Research group consensus</u>: Using second level | NA |
|  | d7701 Spousal relationships | Remove | <u>Research group consensus</u>: Using second level | NA |
|  | d7702 Sexual relationships | Remove | <u>Research group consensus</u>: Using second level | NA |
| **First** | **d8 Major life areas** |  |  |  |
| **Second** | **d850 Remunerative employment** | Revise/<br>add | <u>Think-aloud</u>: Changed to "work".<br>**Quote 41**: "Ohh what is remunerative. What does that mean? "<br><u>Think-aloud</u>: Split into two questions: paid job and unpaid work (adding d855 Non-remunerative employment).<br>**Quote 42**: "depends on what kind of work you're wanting me to do. If you were wanting me to bake and I have to stand in the kitchen to make Christmas dinner; yeah, that's a lot of difficulty. but it depends on what your work is, right? We've got people that have had hip surgery and they worked in construction. So for them, their work construction, that would be horrible for me. I do mine on my computer and I'm sitting. That's not so hard. Paid work is what it should say." | Paid job (part-time or full-time)<br>Unpaid work (such as volunteer work) |
| **Third** | d8500 Self employment | Remove | <u>Think-aloud</u>: Using second level<br>**Quote 43**: "whether it's part-time independent or full time, it's all work, right? It's the same thing. I'm not sure what you're going to get from these questions like it depends on what your job is, what's the purpose of these questions?" | NA |
|  | d8501 Part-time employment | Remove | <u>Think-aloud</u>: Using second level. See **quote 43**. | NA |
|  | d8502 Full-time employment | Remove | <u>Think-aloud</u>: Using second level. See **quote 43**. | NA |
| **First** | **d9 Community, social and civic life** |  |  |  |
| **Second** | **d910 Community Life** | revise | <u>Think-aloud</u>: Merged with "d9205 Socializing" and examples were added from the ICF description.<br>**Quote 44**: "engaging in community such as social. What's important? I come from a big family. We have a lot of relatives and visitors. They come visit me right now. I haven't been going out. " | Socializing /Participate in community life<br>Such as: visiting friends or relatives, meeting in public places, local groups, social organizations, ceremonies (e.g marriages, etc.) |
| **Third** | d9100 Informal associations | Remove | <u>Research group consensus</u>: Using second level | NA |
|  | d9101 Formal associations | Remove | <u>Research group consensus</u>: Using second level | NA |
|  | d9102 Ceremonies | Remove | <u>Research group consensus</u>: Using second level | NA |
| **Second** | **d920 Recreation and leisure** | Remove | Using third level. | NA |
| **Third** | d9200 Play | Revise | <u>Research group consensus:</u> Merged with "d9202 Arts and culture", "d9203 Crafts", and "d9204 Hobbies". Examples were added from the ICF description. | Hobbies/ Play /Crafts/ Arts and culture<br>Such as: gardening, going to the theatre, cinema, museum, reading, singing, playing an instrument, pottery or knitting, playing chess or cards, etc. |
|  | d9201 Sports | Revise | <u>Think-aloud:</u> Added examples of sports common among people undergoing hip or knee arthroplasty.<br>**Quote 34:** *"This would be a bit of a challenge. It would to me it is important to engage in some kind of a sports, especially weight training, would you consider that to be a sport, weight training?"* | Sports<br>such as: Swimming, Golf, Dancing, Workout/gym, Hiking, Skating, Skiing, Bowling, Etc. |
|  | d9202 Arts and culture | Revise | <u>Research group consensus:</u> Merged with " d9200 Play", "d9203 Crafts", and "d9204 Hobbies". Examples were added from the ICF description. | Hobbies/ Play /Crafts/ Arts and culture<br>Such as: gardening, going to the theatre, cinema, museum, reading, singing, playing an instrument, pottery or knitting, playing chess or cards, etc. |
|  | d9203 Crafts | Revise | <u>Research group consensus:</u> Merged with "d9202 Arts and culture", "d9200 Play", and "d9204 Hobbies". Examples were added from the ICF description. | Hobbies/ Play /Crafts/ Arts and culture<br>Such as: gardening, going to the theatre, cinema, museum, reading, singing, playing an instrument, pottery or knitting, playing chess or cards, etc. |
|  | d9204 Hobbies | Revise | <u>Research group consensus:</u> Merged with "d9202 Arts and culture", "d9203 Crafts", and "d9200 Play". Examples were added from the ICF description. | Hobbies/ Play /Crafts/ Arts and culture<br>Such as: gardening, going to the theatre, cinema, museum, reading, singing, playing an instrument, pottery or knitting, playing chess or cards, etc. |
|  | d9205 Socializing | Revise | <u>Research group consensus:</u> Merged with "d910 Community Life". Examples were added from the ICF description. | Socializing /Participate in community life<br>Such as: gardening, going to the theatre, cinema, museum, reading, singing, playing an instrument, pottery or knitting, playing chess or cards, etc. |

### Theme 1: Clarity and comprehension

#### General concepts

All ICF activity descriptions were removed, except those that required further clarification. This approach streamlined the questionnaire by eliminating unnecessary elements to enhance its simplicity and user-friendliness, which can increase the response rate.

The ability to perform activities with or without the use of aids was unclear to some participants. The following clarification was added to the instructions: “Consider your current ability with or without an aid (such as cane, crutches, knee braces, etc.)”.

#### Changing and maintaining body position

The term “maintaining” was replaced with “holding” in all relevant questions, as the former was not clear during the think-aloud interview (*Quote 11*). Since some participants encountered issues related to holding, “how difficult is it to hold…” was changed to “for how long can you hold…”. “Lying down and getting up” and “kneeling down” were unclear for some patients, as the ability could depend on the height of the surface (*Quotes 1,3)*. “In the bed” was added for “lying down and getting up,” and “on the ground” for “kneeling down”. Similarly, the term “partial or full squat” was added to squatting and holding a squatting position (*Quotes 2, 14)*. Some participants found the question about sitting and standing unclear (*Quotes 4-7*). The terms “average chair” and “with or without arms” were added for further clarity. “Such as standing at the sink or in a line” was added to holding a standing position to provide clarification.

“Over or to the side (such as reaching down for an object)” was added to bending (*Quote 8*). “From side to side” was added to shifting the body with the explanation” for example, moving from one foot to another while standing or shifting your weight while sitting” (*Quote 9*). “Such as back to side” was added to rolling over (*Quote 10*). “During the day” was added to holding a lying position for further clarification (*Quotes 12,13*).

#### Lifting and carrying objects

There were issues about the location and weight of the object during lifting and putting down activities (*Quotes 15, 16, 20*). “From the floor” was added to lifting and putting down and divided into light and heavy objects. Also, “less than 1 pound” was used for light and “more than 10 pounds” for heavy.^51,52^ A similar issue was raised for carrying in the hand (*Quote 17*), so “such as a suitcase or grocery bag” was added. The term “hip” in carrying on the shoulder, hip and back was omitted (*Quote 18*), with “such as a backpack” added for clarification (*Quote 19*).

#### Walking

Some participants mentioned that walking short and long distances were unclear (*Quotes 21, 22*), so “less than 1 km” for short distances and “more than 1 km” for long distances were added based on the ICF descriptions. “On flat surfaces” was also added *(Quote 23)*. Additionally, the questions were restructured from “how difficult is it to walk short and long distances” to “How far can you walk short and long distances?”. We excluded the term “ice” as an example in the question concerning walking on different surfaces since participants mentioned it was much more challenging (*Quote 24*).

#### Moving around

The question regarding crawling was removed due to its lack of face validity for patients after arthroplasty (*Quote 25*). Similarly, the terms “ladder” and “rocks” were excluded from the question related to climbing (*Quote 26*). Going up and down stairs was divided into two separate questions, since patients noted that the ability to ascend stairs differs from the ability to descend (*Quote 27*) and “10 to 12 stairs” was added for more clarification (*Quote 28*). Moreover, the term “jogging” was incorporated within the running activity (*Quote 29*). Similarly, “swimming” was moved from “moving around” section to become a sports example (*Quote 30*)

#### Moving around using transportation

Due to ambiguity related to “getting in and out of a car”, “using public transportation”, and “biking” (*Quotes 31-34*), “own car or a taxi”, “a bus or train”, and “to go to a place or for recreation” were added to these activities respectively.

#### Washing oneself

Washing body parts was reworded to “washing lower body parts” to make it more relevant to patients after hip or knee arthroplasty (*Quote 35*).

#### Toileting

Due to ambiguity in the question about going to the toilet (*Quote 36*), we included the description “such as getting to the washroom, getting into and out of position, manipulating clothing, and cleaning oneself” based on the wording from the ICF description.

#### Dressing

The description of putting on and taking off footwear was enriched by incorporating the example of “shoes and/or socks” (*Quote 37*).

Domestic life and intimate relationships

A participant pointed out that shopping could encompass online shopping, which was not aligned with the intended focus (*Quote 38*) and was therefore modified to “going shopping”. The term “intimate” was changed to “sexual” for more clarification (*Quote 40*).

#### Assisting others

The explanation “with their learning, communicating” was excluded (*Quote 39*) to focus on the physical aspect of assisting others.

#### Employment

Participants noted that the abilities to do the activities for self-employment, part-time and full-time employment were similar (*Quote 43*). Consequently, they were unified under the term “work” (*Quote 41*). In consideration of ambiguity related to “work” (*Quote 42*), it was split into two questions: paid job and unpaid work. Furthermore, recognizing that a significant number of patients undergoing hip or knee arthroplasty are retired, we included the response option “I am retired” in the question related to employment.

#### Community, social and civic life

Community life was merged with socializing due to their overlapping concepts (*Quote 44*). Participants stated that the term “sport” was somewhat ambiguous (*Quote 45*). To address this, we provided examples of sports common among people undergoing hip or knee arthroplasty.^53,54^ Moreover, based on existing literature highlighting the impact of dancing on physical health,^55,56^ dancing was reclassified from an art and culture activity to a sport activity.

### Theme 2: Items ordering

Some participants noted challenges in differentiating between changing and maintaining body position. To address this, changing and maintaining body position questions were put one after the other for each activity. There were comments on the order of ability and importance, and importance was put before ability.

We also moved similar activities together, “rolling over” was placed after “lying in bed and getting up”, “putting down objects” was placed after “lifting objects”, “running/jogging” was moved from the “moving around” section to “community, social, and civic life” section after sports.

### Theme 3: Visual elements

Several comments were received regarding the images during the cognitive interviewing process. For example, the picture for walking long distances on flat surfaces seemed like walking on hills. The image depicting walking long distances on flat surfaces was adjusted to more accurately match the question. A research team member (MK) made the following image modifications: “holding the squatting position” from full to partial squat; “walking for long distances” from hills to flat surfaces; “washing the whole body” from sitting to standing; “washing body part” from hands to feet; “standing” from standing from the floor to standing from a chair”; “kneeling” from sitting to kneel to standing to kneel; “lying down” from the floor to bed; “maintaining a sitting position” from a table to a chair. Images for descending stairs, ascending a curb or step, washing lower body parts and rolling over were created by modifying existing ICF illustrations.

## Pilot testing

Following the think-aloud session, we conducted a pilot test of the online questionnaire. This test involved 10 participants, including 5 who underwent hip arthroplasty and 5 who underwent knee arthroplasty within 1, 6, 13, and 26 weeks prior to the study. The primary objective of this initial testing was to assess whether participants encountered any challenges while completing the questionnaire. Participants were free to complete the questionnaire using a computer, tablet or phone.

We administered the questionnaire through Research Electronic Data Capture (REDCap), a secure web application designed for building and managing online surveys and databases for research studies. One of our research team members (MK) converted the questionnaire to the REDCap platform. One researcher (MK) contacted eligible individuals via phone to invite them to participate. We applied an implied consent format placed at the beginning of the questionnaire. During the pilot testing phase, the participants did not face any specific issues or barriers. Minor adjustments were made to the questionnaire design to enhance readability, primarily focused on modifying font styles and sizes to ensure optimal clarity and accessibility for participants.

## Discussion

We developed a questionnaire to determine the activities deemed important by patients with OA undergoing hip or knee arthroplasty based on the activity and participation components of ICF core set for OA. Additionally, we evaluated the face and content validity of the questionnaire through the research team’s refinement and patient’s think-aloud interviews.

The findings of this study indicate that the third-level categories of the OA ICF core set can be used to measure functioning following hip or knee arthroplasty. However, several activities lacked face validity for multiple reasons, including not being relevant to lower extremities, almost never performed, not encompassing all genders, or not being applicable to developed countries, which suggests the need for reconsideration regarding their inclusion for hip or knee arthroplasty. These results align with previous studies suggesting the removal of fine hand use (d440) and hand and arm use (d445) for patients undergoing hip or knee arthroplasty.^28^ It is noteworthy that all previous assessments of the ICF core set for OA ^57–60^ were conducted using the second level categories/items. Our study was the first to use the third level of the OA core set which captures a more comprehensive understanding of patients’ functioning in various aspects of daily life. Moreover, some third level activities within six categories may not be necessary since they require similar abilities justifying the use of second-level categories.

Including all activities adds unnecessary complexity and burden to the evaluation process.

The results of this study suggest that the OA and total knee arthroplasty ICF core sets would benefit from review and modifications to capture the functioning of patients post hip or knee arthroplasty. Previous research has also suggested that the OA core set was missing other relevant categories for patients after arthroplasty.^28^ We recommend adding unpaid work and going up and down stairs to the OA core set, and also adding changing and maintaining body position, driving, shopping, doing housework, assisting others, intimate relationship and community life to the total knee arthroplasty core set. We also recommend adding weight parameters to lifting, adding flat surfaces to walking short and long distances, narrowing the focus of washing to lower body parts, and separating the activities lying down and getting up.

Another issue patients encountered during the think-aloud interviews was the complex wording found in the ICF. The ICF and corresponding core sets were not developed to be completed by patients. It has been argued that measurement instruments should typically be created for a reading level equivalent to age 12.^61^ Hence, we simplified the wording of the ICF framework to make the information accessible to individuals from diverse backgrounds and reading levels.

### Limitations

The study participants are residents of one Canadian province, and the findings may not be directly applicable to other populations. In addition, our recruitment was limited to English-speaking patients who underwent primary elective knee and hip arthroplasty for OA, which may impact the generalizability of our findings. Another limitation was that content analysis of transcripts was performed by one researcher, albeit with peer checking conducted by other research team members. Our recruitment focused on patients at 2, 6, 13, and 26 weeks after hip or knee arthroplasty, limiting the applicability of the questionnaire to other timeframes.

However, since functional recovery mainly takes place during the first 6 months after arthroplasty,^42,43^ it is less likely that the findings would significantly change beyond this timeframe.

## Conclusion

The developed questionnaire based on the ICF core set for OA can be used to assess which activities are important for patients with OA undergoing arthroplasty, contributing to a better understanding of desired outcomes and patient expectations at different time points during post-operative recovery. The questionnaire validation process through the think-aloud interviews identified and addressed issues concerning comprehension, item order, and visual elements.

Furthermore, the pilot study highlighted the feasibility of the online questionnaire and using third-level activities of the ICF core set for OA to assess activity importance for patients after hip or knee arthroplasty.

## Data Availability

All data produced in the present study are available upon reasonable request to the authors

## Acknowledgements

The authors express their gratitude to all the study participants for their time, patience and contributions.

