## Appendix A for "Development and content validation of a questionnaire identifying patients’ functional priorities and abilities after hip or knee arthroplasty"

**Appendix A**. Final version of the questionnaire

### **Please read these important instructions:**

Please rate each activity on two aspects:

1. How **important** the activity is to you **today**. When rating importance, please do not think about your ability to do it. Only consider how important it is for you **today**, not what you think will be important in the future.

2. Your **ability** to do the activity **today**. If you are using an aid (such as a cane, crutches, walker, etc.), assess your ability with the aid you use.

Thank you for participating in our study!

#### Changing and holding a body position

| | | |
|---|---|---|
| **Lying down in bed and getting up**<br>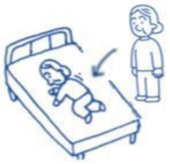<br>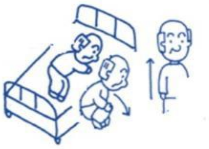 | **Today**, being able to lie down in bed and get up is ... | ☐ Not important<br>☐ Slightly Important<br>☐ Important<br>☐ Very important |
| | **Today**, how difficult is it to lie down in bed and get up? | ☐ Unable to do<br>☐ A lot of difficulty<br>☐ A little difficulty<br>☐ No difficulty<br>☐ I don't know<br>☐ I never do this |
| **Rolling over (such as back to side)**<br>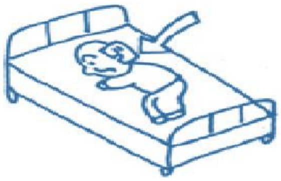 | **Today**, being able to roll over is ... | ☐ Not important<br>☐ Slightly Important<br>☐ Important<br>☐ Very important |
| | **Today**, how difficult is it to roll over? | ☐ Unable to do<br>☐ A lot of difficulty<br>☐ A little difficulty<br>☐ No difficulty<br>☐ I don't know<br>☐ I never do this |
| **Hold a lying position during the day**<br>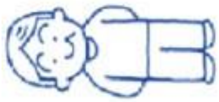 | **Today**, being able to hold a lying position during the day is ... | ☐ Not important<br>☐ Slightly Important<br>☐ Important<br>☐ Very important |
| | **Today**, how long can you hold a lying position during the day? | ☐ Unable to do<br>☐ Less than 10 minutes<br>☐ 11-30 minutes<br>☐ 31-60 minutes<br>☐ More than 1 hour<br>☐ I don't know<br>☐ I never do this |

| | | |
|---|---|---|
| **Squatting down (partial or full squat)**<br>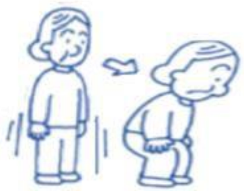 | **<u>Today,</u>** being able to squat down is ... | ☐ Not important<br>☐ Slightly Important<br>☐ Important<br>☐ Very important |
| | **<u>Today</u>**, how difficult is it to squat down? | ☐ Unable to do<br>☐ A lot of difficulty<br>☐ A little difficulty<br>☐ No difficulty<br>☐ I don't know<br>☐ I never do this |
| **Hold a squatting position (partial or full squat)**<br>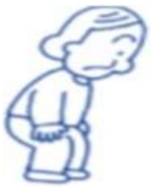 | **<u>Today,</u>** being able to hold a squatting position is ... | ☐ Not important<br>☐ Slightly Important<br>☐ Important<br>☐ Very important |
| | **<u>Today,</u>** how long can you hold a squatting position? | ☐ Unable to do<br>☐ Less than 1 minute<br>☐ 1-5 minutes<br>☐ More than 5 minutes<br>☐ I don't know<br>☐ I never do this |
| **Kneeling down on the ground**<br>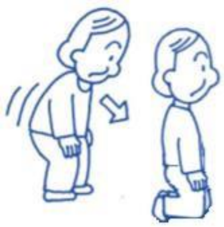 | **<u>Today,</u>** being able to kneel down is ... | ☐ Not important<br>☐ Slightly Important<br>☐ Important<br>☐ Very important |
| | **<u>Today</u>**, how difficult is it to kneel down? | ☐ Unable to do<br>☐ A lot of difficulty<br>☐ A little difficulty<br>☐ No difficulty<br>☐ I don't know<br>☐ I never do this |
| **Hold a kneeling position**<br>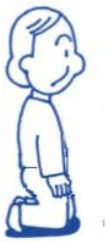 | **<u>Today</u>**, being able to hold a kneeling position is ... | ☐ Not important<br>☐ Slightly Important<br>☐ Important<br>☐ Very important |
| | **<u>Today</u>**, how long can you hold a kneeling position? | ☐ Unable to do<br>☐ Less than 1 minute<br>☐ 1-5 minutes<br>☐ More than 5 minutes<br>☐ I don't know<br>☐ I never do this |

| | | |
|---|---|---|
| **Sitting on a chair (average height with or without arms)**<br>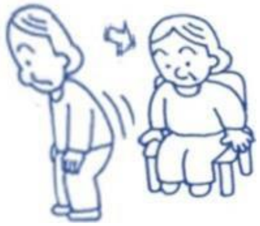 | **<u>Today</u>**, being able to sit on a chair is … | ☐ Not important<br>☐ Slightly Important<br>☐ Important<br>☐ Very important |
| | **<u>Today</u>**, how difficult is it to sit on a chair? | ☐ Unable to do<br>☐ A lot of difficulty<br>☐ A little difficulty<br>☐ No difficulty<br>☐ I don’t know<br>☐ I never do this |
| **Hold a sitting position**<br>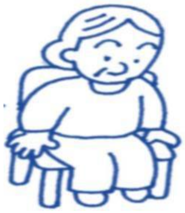 | **<u>Today</u>**, being able to hold a sitting position is … | ☐ Not important<br>☐ Slightly Important<br>☐ Important<br>☐ Very important |
| | **<u>Today,</u>** how long can you hold a sitting position? | ☐ Unable to do<br>☐ Less than 10 minutes<br>☐ 11-30 minutes<br>☐ 31-60 minutes<br>☐ More than 1 hour<br>☐ I don’t know<br>☐ I never do this |
| **Rising from a chair (average height with or without arms)**<br>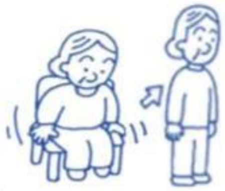 | **<u>Today</u>**, being able to rise from a chair is … | ☐ Not important<br>☐ Slightly Important<br>☐ Important<br>☐ Very important |
| | **<u>Today</u>**, how difficult is it to rise from a chair? | ☐ Unable to do<br>☐ A lot of difficulty<br>☐ A little difficulty<br>☐ No difficulty<br>☐ I don’t know<br>☐ I never do this |
| **Hold a standing position** (such as standing at the sink or in a line)<br>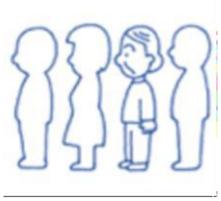 | **<u>Today</u>**, being able to hold a standing position is … | ☐ Not important<br>☐ Slightly Important<br>☐ Important<br>☐ Very important |
| | **<u>Today</u>**, how long can you hold a standing position? | ☐ Unable to do<br>☐ Less than 10 minutes<br>☐ 11-30 minutes<br>☐ More than 30 minutes<br>☐ I don’t know<br>☐ I never do this |

| | | |
|---|---|---|
| **Bending over or to the side**<br>(such as reaching down for an object)<br>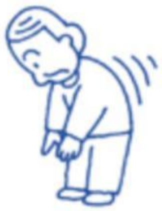 | **<u>Today</u>**, being able to bend over or to the side is ... | ☐ Not important<br>☐ Slightly Important<br>☐ Important<br>☐ Very important |
| | **<u>Today</u>**, how difficult is it to bend over or to the side? | ☐ Unable to do<br>☐ A lot of difficulty<br>☐ A little difficulty<br>☐ No difficulty<br>☐ I don't know<br>☐ I never do this |
| **Shifting the body from side to side**<br>(such as moving from one foot to another while standing or shifting your weight while sitting)<br>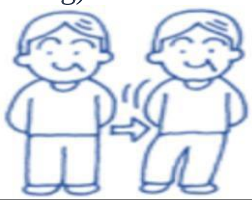 | **<u>Today,</u>** being able to shift your body is ... | ☐ Not important<br>☐ Slightly Important<br>☐ Important<br>☐ Very important |
| | **<u>Today</u>**, how difficult is it to shift your body? | ☐ Unable to do<br>☐ A lot of difficulty<br>☐ A little difficulty<br>☐ No difficulty<br>☐ I don't know<br>☐ I never do this |
| ***Lifting and carrying objects*** | | |
| **Lifting a light object from the floor (less than 1 pound)**<br>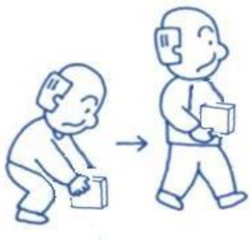 | **<u>Today</u>**, being able to lift a light object from the floor is ... | ☐ Not important<br>☐ Slightly Important<br>☐ Important<br>☐ Very important |
| | **<u>Today</u>**, how difficult is it to lift a light object from the floor? | ☐ Unable to do<br>☐ A lot of difficulty<br>☐ A little difficulty<br>☐ No difficulty<br>☐ I don't know<br>☐ I never do this |
| **Lifting a heavy object from the floor (more than 10 pounds)**<br>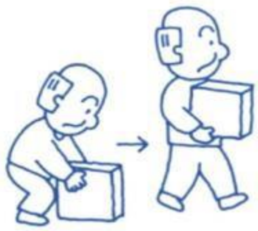 | **<u>Today</u>**, being able to lift a heavy object from the floor is ... | ☐ Not important<br>☐ Slightly Important<br>☐ Important<br>☐ Very important |
| | **<u>Today</u>**, how difficult is it to lift a heavy object from the floor? | ☐ Unable to do<br>☐ A lot of difficulty<br>☐ A little difficulty<br>☐ No difficulty<br>☐ I don't know<br>☐ I never do this |

| | | |
|---|---|---|
| **Putting down objects on the floor**<br>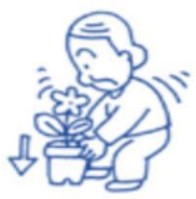 | **<u>Today</u>**, being able to put down objects on the floor is ... | ☐ Not important<br>☐ Slightly Important<br>☐ Important<br>☐ Very important |
| | **<u>Today</u>**, how difficult is it to put down objects on the floor? | ☐ Unable to do<br>☐ A lot of difficulty<br>☐ A little difficulty<br>☐ No difficulty<br>☐ I don't know<br>☐ I never do this |
| **Carrying in the hands**<br>(such as a grocery bag or suitcase)<br>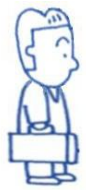 | **<u>Today</u>**, being able to carry an object in your hands is ... | ☐ Not important<br>☐ Slightly Important<br>☐ Important<br>☐ Very important |
| | **<u>Today</u>**, how difficult is it to carry an object in your hands? | ☐ Unable to do<br>☐ A lot of difficulty<br>☐ A little difficulty<br>☐ No difficulty<br>☐ I don't know<br>☐ I never do this |
| **Carrying in the arms**<br>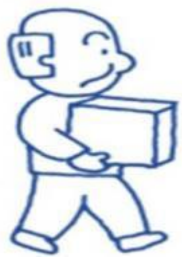 | **<u>Today</u>**, being able to carry an object in your arms is ... | ☐ Not important<br>☐ Slightly Important<br>☐ Important<br>☐ Very important |
| | **<u>Today</u>**, how difficult is it to carry an object in your arms? | ☐ Unable to do<br>☐ A lot of difficulty<br>☐ A little difficulty<br>☐ No difficulty<br>☐ I don't know<br>☐ I never do this |
| **Carrying on shoulders or back**<br>(such as a backpack)<br>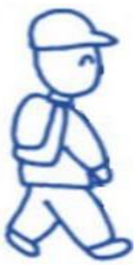 | **<u>Today</u>**, being able to carry an object on your shoulders or back is ... | ☐ Not important<br>☐ Slightly Important<br>☐ Important<br>☐ Very important |
| | **<u>Today</u>**, how difficult is it to carry an object on your shoulders or back? | ☐ Unable to do<br>☐ A lot of difficulty<br>☐ A little difficulty<br>☐ No difficulty<br>☐ I don't know<br>☐ I never do this |

| ***Walking*** | | |
|---|---|---|
| **Walking short distances (less than 1km) on flat surfaces**<br>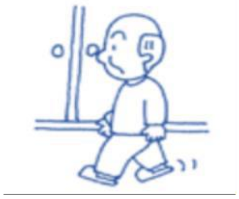 | **<u>Today</u>**, being able to walk short distances is … | ☐ Not important<br>☐ Slightly Important<br>☐ Important<br>☐ Very important |
| | **<u>Today</u>**, how far can you walk short distances (less than 1km)? | ☐ Unable to do<br>☐ Less than 70 steps (50m)<br>☐ 71 - 300 steps (51-200m)<br>☐ 301 - 500 steps (201-500m)<br>☐ 501 - 1500 steps (501m-1km)<br>☐ I don't know<br>☐ I never do this |
| **Walking long distances (more than 1km) on flat surfaces**<br>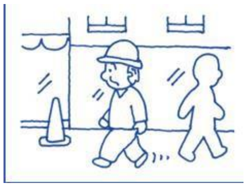 | **<u>Today</u>**, being able to walk long distances is … | ☐ Not important<br>☐ Slightly Important<br>☐ Important<br>☐ Very important |
| | **<u>Today</u>**, how far can you walk long distances (more than 1km)? | ☐ Unable to do<br>☐ 1500 - 5000 steps (1km - 4km)<br>☐ 5001 - 10 000 steps (4 -7km)<br>☐ More than 10 000 steps (more than 7km)<br>☐ I don't know<br>☐ I never do this |
| **Walking on different surfaces** (slope, uneven, or moving surfaces, such as grass, gravel and snow)<br>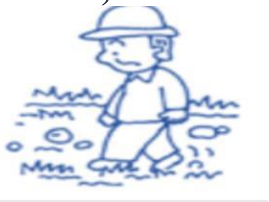 | **<u>Today</u>**, being able to walk on different surfaces is … | ☐ Not important<br>☐ Slightly Important<br>☐ Important<br>☐ Very important |
| | **<u>Today</u>**, how difficult is it to walk on different surfaces? | ☐ Unable to do<br>☐ A lot of difficulty<br>☐ A little difficulty<br>☐ No difficulty<br>☐ I don't know<br>☐ I never do this |
| **Walking around obstacles** (such as walking around a marketplace or shop, around or through crowded areas)<br>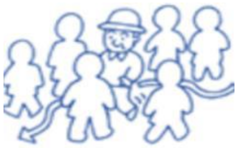 | **<u>Today</u>**, being able to walk around obstacles is … | ☐ Not important<br>☐ Slightly Important<br>☐ Important<br>☐ Very important |
| | **<u>Today</u>**, how difficult is it to walk around obstacles? | ☐ Unable to do<br>☐ A lot of difficulty<br>☐ A little difficulty<br>☐ No difficulty<br>☐ I don't know<br>☐ I never do this |

| Moving around | | |
| --- | --- | --- |
| **Going up stairs (10 to 12 stairs)**<br>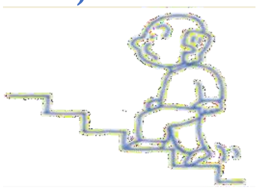 | **<u>Today</u>**, being able to go up stairs is ... | ☐ Not important<br>☐ Slightly Important<br>☐ Important<br>☐ Very important |
| | **<u>Today</u>**, how difficult is it to go up stairs? | ☐ Unable to do<br>☐ A lot of difficulty<br>☐ A little difficulty<br>☐ No difficulty<br>☐ I don't know<br>☐ I never do this |
| **Going down stairs (10 to 12 stairs)**<br>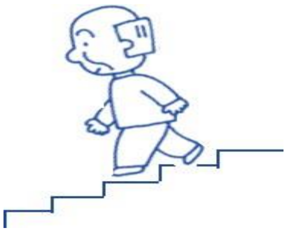 | **<u>Today</u>**, being able to go down stairs is ... | ☐ Not important<br>☐ Slightly Important<br>☐ Important<br>☐ Very important |
| | **<u>Today</u>**, how difficult is it to go down stairs? | ☐ Unable to do<br>☐ A lot of difficulty<br>☐ A little difficulty<br>☐ No difficulty<br>☐ I don't know<br>☐ I never do this |
| **Going up a curb or step**<br>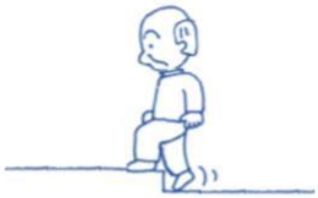 | **<u>Today</u>**, being able to go up a curb or step is ... | ☐ Not important<br>☐ Slightly Important<br>☐ Important<br>☐ Very important |
| | **<u>Today</u>**, how difficult is it to go up a curb or step? | ☐ Unable to do<br>☐ A lot of difficulty<br>☐ A little difficulty<br>☐ No difficulty<br>☐ I don't know<br>☐ I never do this |
| **Jumping**<br>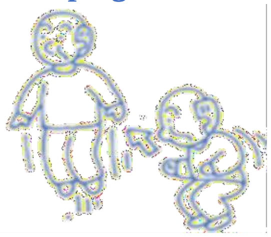 | **<u>Today</u>**, being able to jump is ... | ☐ Not important<br>☐ Slightly Important<br>☐ Important<br>☐ Very important |
| | **<u>Today</u>**, how difficult is it to jump? | ☐ Unable to do<br>☐ A lot of difficulty<br>☐ A little difficulty<br>☐ No difficulty<br>☐ I don't know<br>☐ I never do this |

| ***Moving around using transportation*** | | |
|---|---|---|
| **Getting in and out of a car**<br>(such as own car or taxi)<br>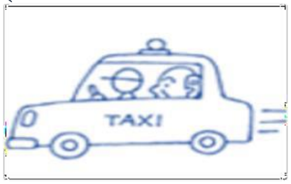 | **<u>Today</u>**, being able to get in and out of a car is … | ☐Not important<br>☐Slightly Important<br>☐Important<br>☐Very important |
| | **<u>Today</u>**, how difficult is it to get in and out of a car? | ☐Unable to do<br>☐ A lot of difficulty<br>☐ A little difficulty<br>☐ No difficulty<br>☐I don't know<br>☐I never do this |
| **Using public transportation**<br>(such as a bus or train)<br>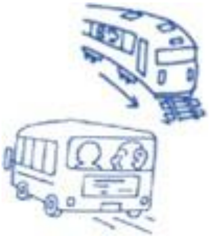 | **<u>Today</u>**, being able to use public transportation is … | ☐Not important<br>☐Slightly Important<br>☐Important<br>☐Very important |
| | **<u>Today</u>**, how difficult is it to use public transportation? | ☐Unable to do<br>☐ A lot of difficulty<br>☐ A little difficulty<br>☐ No difficulty<br>☐ I don't know<br>☐I never do this |
| **Biking (to go to a place or recreational)**<br>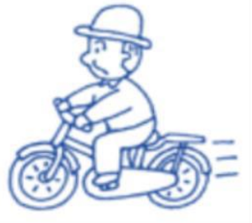 | **<u>Today</u>**, being able to bike is … | ☐Not important<br>☐Slightly Important<br>☐Important<br>☐Very important |
| | **<u>Today</u>**, how difficult is it to bike? | ☐Unable to do<br>☐ A lot of difficulty<br>☐ A little difficulty<br>☐ No difficulty<br>☐ I don't know<br>☐ I never do this |
| **Driving vehicle**<br>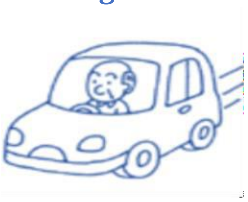 | **<u>Today</u>**, being able to drive vehicle is … | ☐Not important<br>☐Slightly Important<br>☐Important<br>☐Very important |
| | **<u>Today</u>**, how difficult is it to drive vehicle? | ☐Unable to do<br>☐ A lot of difficulty<br>☐A little difficulty<br>☐ No difficulty<br>☐I don't know<br>☐I never do this |

| *Self-care* | | |
|---|---|---|
| **Washing lower body parts**<br>(such as legs and feet)<br>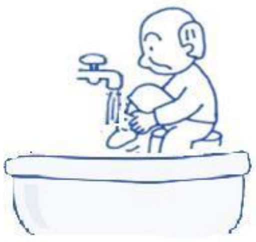 | **<u>Today</u>**, being able to wash your lower body parts is ... | ☐ Not important<br>☐ Slightly Important<br>☐ Important<br>☐ Very important |
| | **<u>Today</u>**, how difficult is it to wash your lower body parts? | ☐ Unable to do<br>☐ A lot of difficulty<br>☐ A little difficulty<br>☐ No difficulty<br>☐ I don't know<br>☐ I never do this |
| **Washing whole body**<br>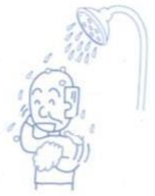 | **<u>Today</u>**, being able to wash your whole body is ... | ☐ Not important<br>☐ Slightly Important<br>☐ Important<br>☐ Very important |
| | **<u>Today</u>**, how difficult is it to wash your whole body? | ☐ Unable to do<br>☐ A lot of difficulty<br>☐ A little difficulty<br>☐ No difficulty<br>☐ I don't know<br>☐ I never do this |
| **Drying your body**<br>(drying some part or parts of your body or the whole body, such as after washing)<br>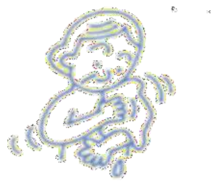 | **<u>Today</u>**, being able to dry your body is ... | ☐ Not important<br>☐ Slightly Important<br>☐ Important<br>☐ Very important |
| | **<u>Today</u>**, how difficult is it to dry your body? | ☐ Unable to do<br>☐ A lot of difficulty<br>☐ A little difficulty<br>☐ No difficulty<br>☐ I don't know<br>☐ I never do this |
| **Going to the toilet**<br>(getting to washroom, getting into and out of position, removing and putting back on clothing, and cleaning oneself.)<br>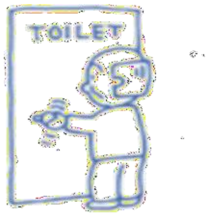 | **<u>Today</u>**, being able to go to the toilet is ... | ☐ Not important<br>☐ Slightly Important<br>☐ Important<br>☐ Very important |
| | **<u>Today</u>**, how difficult is it to go to the toilet? | ☐ Unable to do<br>☐ A lot of difficulty<br>☐ A little difficulty<br>☐ No difficulty<br>☐ I don't know<br>☐ I never do this |

| | | |
|---|---|---|
| **Putting on clothes**<br>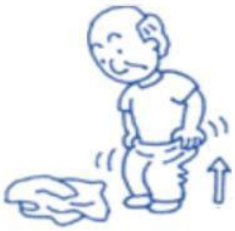 | **Today**, being able to put on clothes is … | ☐ Not important<br>☐ Slightly Important<br>☐ Important<br>☐ Very important |
| | **Today**, how difficult is it to put on clothes? | ☐ Unable to do<br>☐ A lot of difficulty<br>☐ A little difficulty<br>☐ No difficulty<br>☐ I don't know<br>☐ I never do this |
| **Taking off clothes**<br>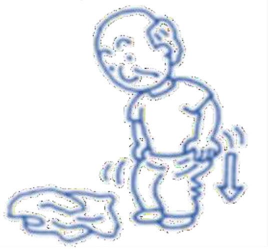 | **Today**, being able to take off clothes is … | ☐ Not important<br>☐ Slightly Important<br>☐ Important<br>☐ Very important |
| | **Today**, how difficult is it to take off clothes? | ☐ Unable to do<br>☐ A lot of difficulty<br>☐ A little difficulty<br>☐ No difficulty<br>☐ I don't know<br>☐ I never do this |
| **Putting on footwear (shoes or socks)**<br>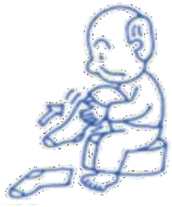 | **Today**, being able to put on footwear is … | ☐ Not important<br>☐ Slightly Important<br>☐ Important<br>☐ Very important |
| | **Today**, how difficult is it to put on footwear? | ☐ Unable to do<br>☐ A lot of difficulty<br>☐ A little difficulty<br>☐ No difficulty<br>☐ I don't know<br>☐ I never do this |
| **Taking off footwear (shoes or socks)**<br>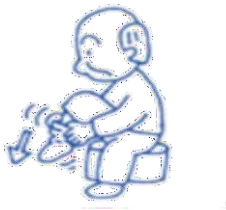 | **Today**, being able to take off footwear is … | ☐ Not important<br>☐ Slightly Important<br>☐ Important<br>☐ Very important |
| | **Today**, how difficult is it to take off footwear? | ☐ Unable to do<br>☐ A lot of difficulty<br>☐ A little difficulty<br>☐ No difficulty<br>☐ I don't know<br>☐ I never do this |

| ***Domestic life and Intimate relationships*** | | |
|---|---|---|
| **Going shopping**<br>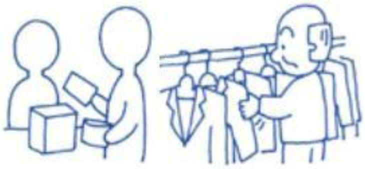 | **Today**, being able to go shopping is ... | ☐ Not important<br>☐ Slightly Important<br>☐ Important<br>☐ Very important |
| | **Today**, how difficult is it to go shopping? | ☐ Unable to do<br>☐ A lot of difficulty<br>☐ A little difficulty<br>☐ No difficulty<br>☐ I don't know<br>☐ I never do this |
| **Doing housework**<br>(cleaning the house, washing clothes, using household appliances, storing food and disposing of garbage)<br>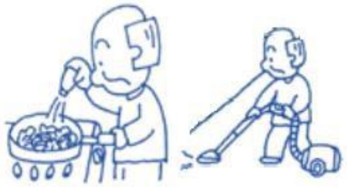 | **Today**, being able to do housework is … | ☐ Not important<br>☐ Slightly Important<br>☐ Important<br>☐ Very important |
| | **Today**, how difficult is it to do housework? | ☐ Unable to do<br>☐ A lot of difficulty<br>☐ A little difficulty<br>☐ No difficulty<br>☐ I don't know<br>☐ I never do this |
| **Assisting others**<br>(assisting others with their self-care, movement, within the house or outside)<br>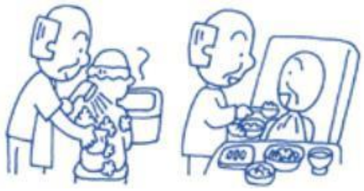 | **Today**, being able to assist others is ... | ☐ Not important<br>☐ Slightly Important<br>☐ Important<br>☐ Very important |
| | **Today**, how difficult is it to assist others? | ☐ Unable to do<br>☐ A lot of difficulty<br>☐ A little difficulty<br>☐ No difficulty<br>☐ I don't know<br>☐ I never do this |
| **Sexual relationships**<br>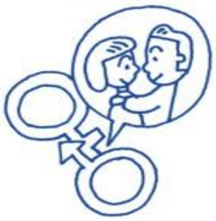 | **Today**, being able to have sexual relationships is ... | ☐ Not important<br>☐ Slightly Important<br>☐ Important<br>☐ Very important |
| | **Today**, how difficult is it to have sexual relationships? | ☐ Unable to do<br>☐ A lot of difficulty<br>☐ A little difficulty<br>☐ No difficulty<br>☐ I don't know<br>☐ I never do this |

| | | |
|---|---|---|
| ***Employment*** | | |
| **Paid job (part-time, or full-time)** 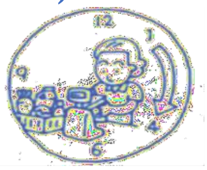 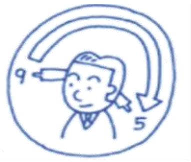 | **<u>Today</u>**, being able to do a paid job is ... | ☐Not important<br>☐Slightly Important<br>☐Important<br>☐Very important |
| | **<u>Today</u>**, how difficult is it to do a paid job? | ☐Unable to do<br>☐ A lot of difficulty<br>☐ A little difficulty<br>☐ No difficulty<br>☐I don't know<br>☐I am retired/ I never do this |
| **Unpaid work (such as volunteer work)** 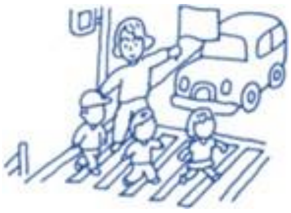 | **<u>Today</u>**, being able to do unpaid work is ... | ☐Not important<br>☐Slightly Important<br>☐Important<br>☐Very important |
| | **<u>Today</u>**, how difficult is it to do unpaid work? | ☐Unable to do<br>☐ A lot of difficulty<br>☐ A little difficulty<br>☐ No difficulty<br>☐ I don't know<br>☐I never do this |
| ***Community, social and civic life*** | | |
| **Socializing /Participate in community life**<br>such as:<br>• Visiting friends or relatives<br>• Meeting in public places, local groups, social organizations, ceremonie s (e.g marriages, etc.) 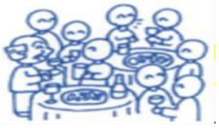 | **<u>Today</u>**, being able to socialize and participate in community life is ... | ☐Not important<br>☐Slightly Important<br>☐Important<br>☐Very important |
| | **<u>Today</u>**, how difficult is it to socialize and participate in community life? | ☐Unable to do<br>☐ A lot of difficulty<br>☐ A little difficulty<br>☐ No difficulty<br>☐I don't know<br>☐I never do this |

| | | |
|---|---|---|
| **Sports**<br>such as:<br>• Swimming<br>• Golf<br>• Dancing<br>• Workout/gym<br>• Hiking<br>• Skating<br>• Skiing<br>• Bowling<br>• Etc.<br>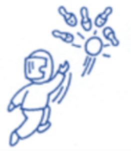 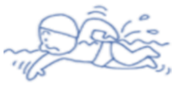 | **Today,** being able to do sports is ... | ☐ Not important<br>☐ Slightly Important<br>☐ Important<br>☐ Very important |
| | **<u>Today</u>**, how difficult is it to do sports? | ☐ Unable to do<br>☐ A lot of difficulty<br>☐ A little difficulty<br>☐ No difficulty<br>☐ I don't know<br>☐ I never do this |
| **Running/jogging**<br>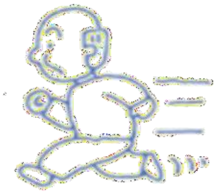 | **<u>Today</u>**, being able to run or jog is ... | ☐ Not important<br>☐ Slightly Important<br>☐ Important<br>☐ Very important |
| | **<u>Today</u>**, how difficult is it to run or jog? | ☐ Unable to do<br>☐ A lot of difficulty<br>☐ A little difficulty<br>☐ No difficulty<br>☐ I don't know<br>☐ I never do this |
| **Hobbies/ play /Crafts/ Arts and culture**<br>Such as:<br>• Gardening<br>• Going to the theatre, cinema, museum<br>• Reading<br>• Singing<br>• Playing an instrument<br>• Pottery or knitting<br>• Playing chess or cards<br>• Etc.<br>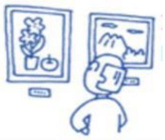 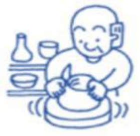 | **<u>Today</u>**, being able to participate in hobbies, play, crafts, arts and culture is ... | ☐ Not important<br>☐ Slightly Important<br>☐ Important<br>☐ Very important |
| | **<u>Today</u>**, how difficult is it to participate in hobbies, play, crafts, arts and culture? | ☐ Unable to do<br>☐ A lot of difficulty<br>☐ A little difficulty<br>☐ No difficulty<br>☐ I don't know<br>☐ I never do this |

| If **<u>other activities</u>** important to you **<u>today</u>** were not included in this questionnaire, please tell us about them. Rate each activity for your ability to do it and its importance **<u>today</u>**. | |
|---|---|
| 1………………… | ☐ Slightly Important<br>☐ Important<br>☐ Very important<br>☐ Unable to do<br>☐ A lot of difficulty<br>☐ A little difficulty<br>☐ No difficulty<br>☐ I don't know<br>☐ I never do this |
| 2…………………. | ☐ Slightly Important<br>☐ Important<br>☐ Very important<br>☐ Unable to do<br>☐ A lot of difficulty<br>☐ A little difficulty<br>☐ No difficulty<br>☐ I don't know<br>☐ I never do this |
| 3…………………. | ☐ Slightly Important<br>☐ Important<br>☐ Very important<br>☐ Unable to do<br>☐ A lot of difficulty<br>☐ A little difficulty<br>☐ No difficulty<br>☐ I don't know<br>☐ I never do this |
| 4…………………………. | ☐ Slightly Important<br>☐ Important<br>☐ Very important<br>☐ Unable to do<br>☐ A lot of difficulty<br>☐ A little difficulty<br>☐ No difficulty<br>☐ I don't know<br>☐ I never do this |

**Please answer the following questions.**

Which surgery did you undergo?
Hip replacement ☐ Knee replacement ☐

Are you currently avoiding certain movements or positions based on your doctor or physiotherapist recommendations?
Yes ☐ No ☐

On average, how much pain have you had in your operated hip or knee in the past 24 hours, 0 being no Worse pain and 100 the worst pain imaginable?

No pain ---------- Moderate pain ---------- Worse pain imaginable

(Place a mark on the scale above)

Do you have any problems or pain in other joints beyond the operated joint?
Yes ☐ No ☐

If yes, the problems or pain is in which joint (s)? Please check all that apply.
Right hip ☐ Right knee ☐ Right foot ☐
Left hip ☐ Left knee ☐ Left foot ☐
Low back ☐ neck ☐ Shoulder ☐

Do you presently use any walking aids? Please check all that apply.

Wheelchair ☐ Walker ☐ Cane ☐ Crutches ☐ Scooter ☐
Other walking aids ☐ Not using any aid ☐

With whom do you live?
alone ☐ with family ☐

Do you have family/relatives that live nearby?
Yes ☐ No ☐

Do you have someone who is dependent on you for their care?
Yes ☐ No ☐

What is your place of residence?
House ☐ Apartment ☐ Retirement Home ☐ Long Term Care Facility ☐ Other ☐

What is the level of activity of your job?

Sedentary ☐ Light physical ☐ Heavy physical ☐ Retired ☐

| | |
|---|---|
| | What is your age? |
| | What was your sex at birth?<br>Male ☐ Female ☐<br><br>What is your gender?<br>Man ☐ Woman ☐ or: please specify ☐ |
| | In order to calculate your body mass index, please provide your height and weight.<br>Height feet/inches<br>Weight lbs |
| | You may belong to one or more racial or cultural groups on the following list. Select all that apply.<br>White ☐<br>First Nation, Métis, Inuk (Inuit) ☐<br>South Asian (e.g., East Indian, Pakistan, Sri Lankan) ☐<br>Chinese ☐<br>Black ☐<br>Filipino ☐<br>Latin American ☐<br>Arab ☐<br>Southeast Asian (e.g., Vietnamese, Cambodian, Malaysian, Laotian) ☐<br>West Asian (e.g., Iranian, Afghan) ☐<br>Korean ☐<br>Japanese ☐<br>Prefer not to say ☐<br>Do not know ☐<br>Other-Specify: |

**Thank you for taking the time to complete this questionnaire!**
